# Building trust through collaboration: A mixed-methods evaluation of San Francisco’s Pregnancy Village model of cross-sector care delivery

**DOI:** 10.1101/2025.08.16.25333840

**Authors:** Osamuedeme J. Odiase, April J. Bell, Alison M. El Ayadi, Catherine Ravikumar, Kattia Vargas, KaSelah Crockett, Malini A. Nijagal, Patience A. Afulani

**Affiliations:** Department of Obstetrics, Gynecology, & Reproductive Sciences, University of California, San Francisco, San Francisco, California, United States; Department of Family and Community Medicine, University of California, San Francisco, California, United States; Department of Epidemiology and Biostatistics, University of California, San Francisco, San Francisco, California, United States; Department of Neurological Surgery, Weill Institute for Sciences, University of California, San Francisco, California, United States; Institute for Health and Aging, University of California, San Francisco, San Francisco, CA, USA; Compass & Keys, Oakland, California, United States; Pop-Up Village, Oakland, California, United States

**Keywords:** trust, medical mistrust, community-institution partnership, health system, community-based organizations, pregnancy, perinatal health, birth equity, health equity, program evaluation

## Abstract

**Background:** Historical injustices, systemic racism, unequal healthcare access, and provider bias have fostered mistrust in healthcare institutions. Cross-sector collaborations between healthcare institutions and community-based organizations (CBOs), such as San Francisco’s Pregnancy Village (PV) model, could potentially build institutional trust in minoritized communities. This study primarily aimed to examine trust in PV, with secondary aims exploring participant perceptions of trust in the health system and CBOs, including their views on the health system’s involvement in PV.

**Methods:** Between July 2021 and June 2022, we conducted a convergent, mixed-methods study involving 116 survey participants (57 pregnant/postpartum individuals and 59 family members) and 18 semi-structured interviews (13 pregnant/postpartum people and five family members). Trust was assessed quantitatively using a seven-item scale (scores standardized to 0-100) and qualitatively with open-ended questions. We performed univariate, bivariate, and multivariate analyses of the quantitative data and thematic analyses of the qualitative data.

**Results:** The mean trust in PV score was 85.9/100 (SD = 18.9). Lower trust was observed among Latine participants, those with a history of preterm birth, and those experiencing food insecurity. Qualitative findings revealed that trust in both the health system and CBOs was shaped by receipt of person-centered care. Trust in CBOs was attributed to their focus on holistic care, relatability, and responsiveness to community needs. Distrust in the health system was shaped by experiences of racism and neglect. Participants held mixed views on the health system’s role in PV—some highlighted its positive contributions, while others voiced skepticism due to ongoing structural racism and inequities in care.

**Conclusions:** Participants perceived PV as trustworthy, with mixed views of the health system, generally positive perceptions of CBOs, and overall support for the health system’s involvement in PV despite lingering concerns. However, concerns about structural racism and healthcare inequities persist. Sustained cross-sector collaboration, guided by community priorities, is critical to building trust and addressing structural inequities.

## BACKGROUND

Historical injustices, such as the Tuskegee Syphilis Study, forced sterilization under state eugenics programs, and medical exploitation of Black women, have fostered an entrenched mistrust of healthcare among Black individuals (1–4). These injustices persist today through structural, systemic, and interpersonal racism, leading to unequal healthcare access and quality, which exacerbate disparities in maternal health (5–8) and reinforce a cycle of negative engagement with healthcare systems (9). Community-based organizations (CBOs)—non-profit, non-governmental, or charitable organizations committed to addressing community needs (10)—have emerged as pivotal stakeholders within the health ecosystem, which encompasses healthcare systems, CBOs, and public health and government agencies. CBOs have taken on an increasingly vital role in response to longstanding deficiencies in traditional healthcare systems (networks of hospitals and clinics that work together to deliver healthcare) (11,12). Often characterized as part of the “third sector,” CBOs occupy a unique intermediary space, filling the gap between state-provided social services and traditional healthcare systems (13,14).

CBOs play a vital role in Black communities, often acting as first responders in efforts to combat disparities. They address critical service gaps in areas like maternal and child health—providing doulas, community health workers, and referrals to other social services—as well as education, housing, and economic empowerment through community-driven, culturally responsive care approaches (15,16). For instance, CBOs like SisterWeb provide free doula support and advocacy for Black families (17); the Homeless Prenatal Program provides housing support and other services to low-income families (18); and Rafiki Coalition for Health and Wellness provides a safe space for therapy, massages, chiropractic services, and other community resources in San Francisco (19). Over time, their role has expanded from meeting urgent and immediate needs to advocating for systemic change, social justice, and health equity in response to pervasive inequities and institutional neglect (21–23). However, organizational barriers, such as perennial underfunding and staffing shortages, limit their ability to scale services in response to community demand (24–26), undermining trust.

CBOs have increasingly collaborated with health systems to share and optimize resources, recognizing that their collaboration could achieve a greater collective impact than either sector could accomplish independently (27). In parallel, health system leaders are engaging CBOs more actively in the development of health programs, services, and interventions (11,28–32). Evidence further underscores that the inclusion of CBOs in decision-making processes enhances the cultural relevance, acceptability, and effectiveness of health interventions (11,33,34), thereby contributing to increased institutional trust (27,35). However, these cross-sector collaborations are not without their challenges, as these collaborations are often hindered by power asymmetries, whereby health systems dominate decision-making and set priorities without adequate community input (36–38). Additionally, CBOs are frequently expected to shoulder the dual responsibility of serving the community and building trust, often devoid of sustained health system-level reforms or institutional support (39).

Given these structural and relational barriers, there is a pressing need to reimagine how these sectors work together to address community needs and build trust. Addressing these challenges, therefore, requires innovative, cross-sector, community-driven models designed to build trust in Black and minoritized communities (40). One such model is the “Pregnancy Village” (PV) model—a cross-sector collaboration, providing a “one-stop shop” of comprehensive wellness services in a safe and uplifting environment for those who face the starkest inequities: Black-identifying pregnant and postpartum individuals. PV aims to build trust and holistically improve the health and wellness of individuals, families, and communities through a person-centered, anti-racist, and culturally grounded approach. Following a three-year planning phase in collaboration with the creators of the “Pop-Up Village” event model, Designing Justice + Designing Spaces, the first iteration of the Pregnancy Village—the “SF Family & Pregnancy Pop-Up Village” (subsequently referred to as the “Pregnancy Village” (PV) for brevity)—was launched as a monthly event series from July 2021 to June 2024 (42). Further details on the planning phase and launch of PV can be found elsewhere (42,43).

Trust is vital for sustaining community sociomedical interventions like PV. Therefore, as part of the evaluation of PV, we sought to assess participants’ perceptions of PV’s trustworthiness and key predictors. Additionally, we explored participants’ perceptions of trust in CBOs and the health system, as well as their perceptions of the health system’s role in PV. This paper presents findings from the formative phase—the first nine months of PV implementation (July 2021-June 2022)—as part of ongoing community engagement efforts to refine the PV model.

## METHODS

### Setting

The Pregnancy Village (PV) holds events in San Francisco, California’s Bayview district, a neighborhood of 35,000 residents, known as the city’s “most isolated” (44), with significant inequities in care access, experience, and outcomes. Most birthing individuals in Bayview are Medicaid-insured (62%), and 93% are from racial or ethnic minority groups (45). Bayview residents also experience markedly lower rates of timely prenatal care, higher rates of preterm birth, and low birth weight (46). The Bayview has industrial, commercial, and residential zones, but declining industry has led to reduced infrastructure investment (47). PV events were held near a major roadway to leverage proximity to public transportation and CBOs, aiming to build trust and foster community engagement (47).

### Intervention

PV is a model for cross-sector collaboration designed to address perinatal inequities by providing a one-stop shop for resources in a celebratory and uplifting environment for Black pregnant individuals and their families. This care delivery model is rooted in person-centered and anti-racism principles, promoting sustainable community-institutional partnerships and featuring a real-time community feedback system to ensure responsive model iteration based on community’s stated needs (48). PV features six distinct resource hubs (neighborhoods), each representing essential components needed to support comprehensive wellness within communities (42) (Figure 1). The event design features vibrant colors, shaded areas, ground treatments, and varied seating arrangements to create an inviting space. Further details about the implementation of PV can be found elsewhere (42).

**Figure 1.**
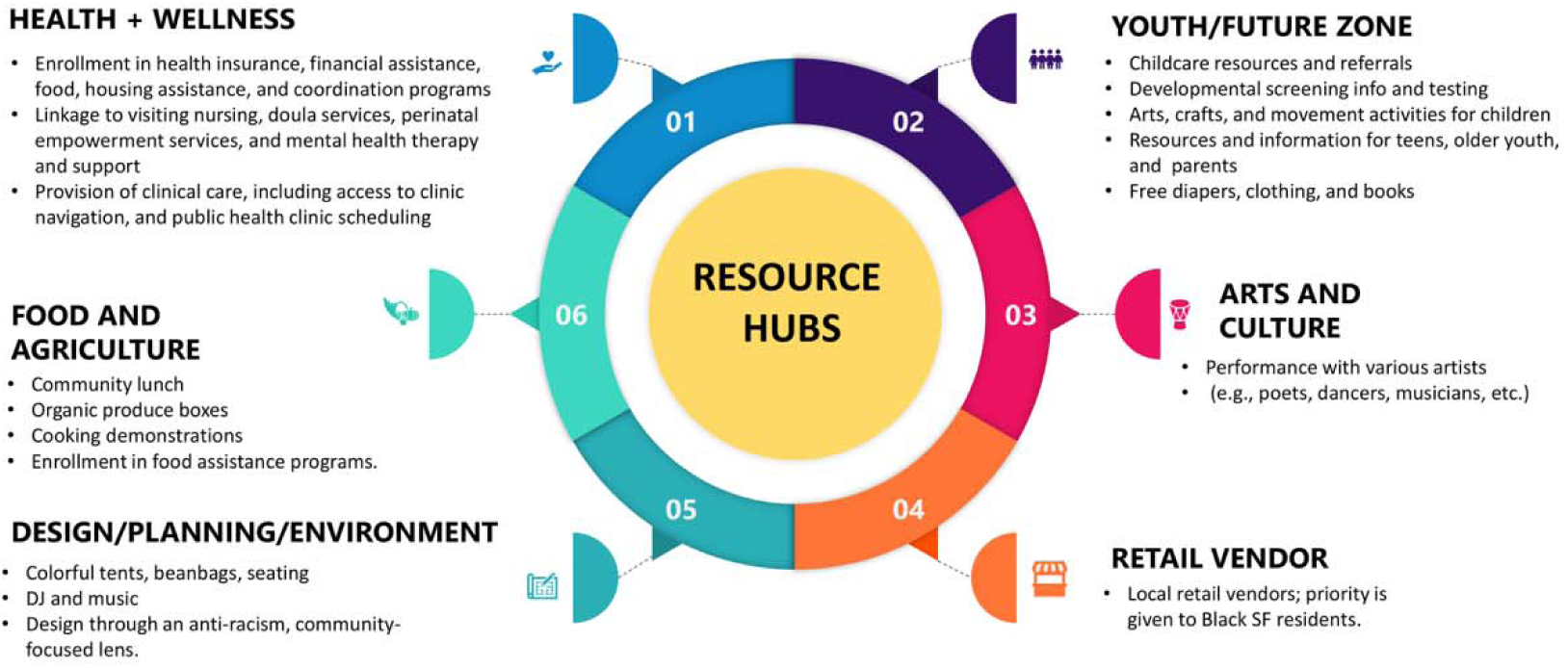
Family and Pregnancy Pop-Up Village resource hubs and summary of provided services. This figure depicts the core elements of the Pregnancy Village model, which aims to provide a one-stop shop of cross-sector services in an uplifting, safe, and supportive environment for pregnant individuals and their families.

### Study design

We conducted a convergent mixed-methods, community-engaged evaluation, in which quantitative and qualitative data were collected concurrently through surveys and in-depth reviews, analyzed independently, and then integrated to interpret the findings. Integration was further supported by selecting a subset of participants for the in-depth interviews and developing the interview guide to explore topics introduced in the survey. This paper presents data related to trust in PV and participant perceptions of trust in the health system and CBOs, including their views on the health system’s involvement in PV.

### Sample

We recruited a convenience sample of pregnant and postpartum individuals, along with family members participating in the first nine monthly PV events between July 2021 and June 2022. The inclusion criteria were: 1) individuals must be at least 15 years old if pregnant or postpartum, or 18 years old if they were family members; 2) attendance at a minimum of one PV event; and 3) the ability to speak either English or Spanish. We focused on recruiting Black and other minoritized pregnant or postpartum individuals and their families, given that they were the focus population for PV, with a goal of 120 participants (10-15 individuals from each event) for the surveys. We then purposively sampled a subset of 18 participants (13 pregnant and postpartum individuals and 5 family members) for semi-structured in-depth interviews, guided by the principle of maximum variation to capture a wide range of experiences, including race/ethnicity, language, pregnancy status, and age (49). Interviews ended when thematic saturation was achieved (50).

Eligible participants were recruited during PV registration, where study team members provided details about the study and obtained verbal informed consent. Those who consented were given the option to complete a survey onsite using a study tablet or later on their own device via a QR code. Most participants completed the survey onsite and were compensated with a $20 gift card for their participation. Those who attended multiple PV events were permitted to complete the survey more than once.^1^ In addition, three to four participants from each PV event were approached during the events or contacted by phone to invite them for one-on-one in-depth interviews. Interviews were held within four weeks of attending a PV event, scheduled at participants’ convenience, and conducted via Zoom in English or Spanish. The semi-structured interview guide used included questions to explore: 1) trust in the health system; 2) trust in CBOs; and 3) perceptions of the health system’s involvement in PV to help understand trust in PV. Interviews lasted 30-60 minutes and were conducted by trained qualitative researchers (OJO and KV). Interview participants were compensated an additional $20. With participants’ consent, the interviews were recorded, transcribed by a third-party transcriptionist, and verified by study team members (KV and JV). Field notes were taken during the interviews, and a standardized template aided in rapid analysis for model iteration and assessment of saturation (42). These methods have been previously described elsewhere (47).

### Quantitative measures

*Trust in PV*: Trust in PV was evaluated using a 7-item scale adapted from a validated 23-item scale designed to assess trust in the public healthcare system (51) (See individual items in Additional file 1). The adapted scale consisted of seven closed-ended questions, with responses provided on a 4-point Likert scale: 3 for “Strongly agree,” 2 for “Agree,” 1 for “Disagree,” and 0 for “Strongly disagree.” The responses were summed and then standardized on a scale of 0 to 100 by dividing the score by the maximum possible score and multiplying by 100, with higher scores indicating a higher level of trust. Missing data (2.6%) were imputed with the mean of the other responses within the measure. The internal reliability consistency for the trust scale within this sample was Cronbach’s α = 0.95.

*Covariates*: Participants self-reported sociodemographic characteristics, pregnancy and obstetric factors, and experiences of discrimination (See Table 1).

**Table 1.** Characteristics of Pregnancy Village study participants.

|  | Total (N=116) | Currently pregnant/recently pregnant (n=57) | Family member (n=59) |
| --- | --- | --- | --- |
| **Participant Characteristics** | **% (N)** | **% (n)** | **% (n)** |
| *Age* |  |  |  |
| 15-24 | 11.2% (13) | 17.5% (10) | 5.1% (3) |
| 25-34 | 26.7% (31) | 38.6% (22) | 15.3% (9) |
| 35-44 | 21.6% (25) | 33.3% (19) | 10.2% (6) |
| 45 and older | 25.9% (30) | 1.8% (1) | 49.2% (29) |
| Unknown | 14.7% (17) | 8.8% (5) | 20.3% (12) |
| *Gender* |  |  |  |
| Female | 86.2% (100) | 91.2% (52) | 81.4% (48) |
| Male | 5.2% (6) | 0.0% (0) | 10.2% (6) |
| Other/unknown/prefer not to answer | 8.6% (10) | 8.8% (5) | 8.5% (5) |
| *Race/ethnicity* |  |  |  |
| Non-Hispanic Black | 40.5% (47) | 26.3% (15) | 54.2% (32) |
| Hispanic/Latine | 36.2% (42) | 47.4% (27) | 25.4% (15) |
| Multiracial | 15.5% (18) | 19.3% (11) | 11.9% (7) |
| Other/unknown/prefer not to answer | 7.8% (9) | 7.0% (4) | 8.5% (5) |
| *Language* |  |  |  |
| English | 59.5% (69) | 52.6% (30) | 66.1% (39) |
| Spanish | 30.2% (35) | 38.6% (22) | 22.0% (13) |
| Unknown/other/prefer not to answer | 10.3% (12) | 8.8% (5) | 11.9% (7) |
| *English proficiency* |  |  |  |
| Very well or well | 70.7% (82) | 70.2% (40) | 71.2% (42) |
| With difficulty | 17.2% (20) | 19.3% (11) | 15.3% (9) |
| Unknown/prefer not to answer | 12.1% (14) | 10.5% (6) | 13.6% (8) |
| *Pregnancy status* |  |  |  |
| Family member | 50.9% (59) | 0.0% (0) | 100.0% (59) |
| Pregnant or postpartum | 49.1% (57) | 100.0% (57) | 0.0% (0) |
| *Parity* |  |  |  |
| No births | 22.4% (26) | 26.3% (15) | 18.6% (11) |
| 1-2 births | 43.1% (50) | 47.4% (27) | 39.0% (23) |
| 3 or more births | 24.1% (28) | 24.6% (14) | 23.7% (14) |
| Unknown/not applicable/prefer not to answer | 10.3% (12) | 1.8% (1) | 18.6% (11) |
| *Prenatal care* |  |  |  |
| No prenatal care during current/recent pregnancy | 3.4% (4) | 7.0% (4) | 0.0% (0) |
| Received prenatal care during current/recent pregnancy | 41.4% (48) | 84.2% (48) | 0.0% (0) |
| Unknown/not applicable/ prefer not to answer | 55.2% (64) | 8.8% (5) | 100.0% (59) |
| *History of preterm birth* |  |  |  |
| No preterm births | 69.0% (80) | 80.7% (46) | 57.6% (34) |
| At least 1 preterm birth | 19.8% (23) | 15.8% (9) | 23.7% (14) |
| Unknown/not applicable/prefer not to answer | 11.2% (13) | 3.5% (2) | 18.6% (11) |
| *History of pregnancy loss* |  |  |  |
| No prior pregnancy loss | 59.5% (69) | 57.9% (33) | 61.0% (36) |
| Pregnancy loss | 25.0% (29) | 35.1% (20) | 15.3% (9) |
| Unknown/not applicable/prefer not to answer | 15.5% (18) | 7.0% (4) | 23.7% (14) |
| *Education* |  |  |  |
| Less than a high school degree | 18.1% (21) | 15.8% (9) | 20.3% (12) |
| High school graduate, GED or equivalent | 25.9% (30) | 29.8% (17) | 22.0% (13) |
| Some college, junior college or vocational school | 27.6% (32) | 21.1% (12) | 33.9% (20) |
| College graduate or higher | 17.2% (20) | 21.1% (12) | 13.6% (8) |
| Unknown/prefer not to answer | 11.2% (13) | 12.3% (7) | 10.2% (6) |
| *Employment* |  |  |  |
| No | 63.8% (74) | 56.1% (32) | 71.2% (42) |
| Yes, Full-time | 13.8% (16) | 19.3% (11) | 8.5% (5) |
| Yes, Part-time | 10.3% (12) | 15.8% (9) | 5.1% (3) |
| Unknown/prefer not to answer | 12.1% (14) | 8.8% (5) | 15.3% (9) |
| *Income assistance* |  |  |  |
| No | 43.1% (50) | 33.3% (19) | 52.5% (31) |
| Yes | 47.4% (55) | 54.4% (31) | 40.7% (24) |
| Unknown/prefer not to answer | 9.5% (11) | 12.3% (7) | 6.8% (4) |
| *Residence* |  |  |  |
| Bayview-Hunter's Point | 33.6% (39) | 38.6% (22) | 28.8% (17) |
| Other San Francisco | 44.8% (52) | 43.9% (25) | 45.8% (27) |
| East Bay | 9.5% (11) | 12.3% (7) | 6.8% (4) |
| Other/unknown/prefer not to answer | 12.1% (14) | 5.3% (3) | 18.6% (11) |
| *Housing* |  |  |  |
| Homeless Shelter | 10.3% (12) | 12.3% (7) | 8.5% (5) |
| Living with someone for free | 4.3% (5) | 5.3% (3) | 3.4% (2) |
| No living place | 0.9% (1) | 0.0% (0) | 1.7% (1) |
| Owens home or apartment | 6.9% (8) | 5.3% (3) | 8.5% (5) |
| Public Housing | 14.7% (17) | 12.3% (7) | 16.9% (10) |
| Renting home or apartment | 50.0% (58) | 52.6% (30) | 47.5% (28) |
| Transitional or supportive housing | 2.6% (3) | 3.5% (2) | 1.7% (1) |
| Other/unknown/prefer not to answer | 10.3% (12) | 8.8% (5) | 11.9% (7) |
| *Social Support* |  |  |  |
| Not at all | 12.9% (15) | 7.0% (4) | 18.6% (11) |
| A little | 9.5% (11) | 14.0% (8) | 5.1% (3) |
| Somewhat | 19.0% (22) | 22.8% (13) | 15.3% (9) |
| Yes, definitely | 54.3% (63) | 54.4% (31) | 54.2% (32) |
| Unknown/prefer not to answer | 4.3% (5) | 1.8% (1) | 6.8% (4) |
| *Medical insurance status* |  |  |  |
| Public insurance | 66.4% (77) | 66.7% (38) | 66.1% (39) |
| Private insurance | 15.5% (18) | 19.3% (11) | 11.9% (7) |
| No insurance | 4.3% (5) | 1.8% (1) | 6.8% (4) |
| Unknown/prefer not to answer | 13.8% (16) | 12.3% (7) | 15.3% (9) |
| *Food insecurity (Worried food would run out)* |  |  |  |
| Often true | 11.2% (13) | 8.8% (5) | 13.6% (8) |
| Sometimes true | 24.1% (28) | 33.3% (19) | 15.3% (9) |
| Never true | 48.3% (56) | 36.8% (21) | 59.3% (35) |
| Unknown/prefer not to answer | 16.4% (19) | 21.1% (12) | 11.9% (7) |
| *Food insecurity (Insufficient funds)* |  |  |  |
| Often true | 11.2% (13) | 12.3% (7) | 10.2% (6) |
| Sometimes true | 27.6% (32) | 36.8% (21) | 18.6% (11) |
| Never true | 43.1% (50) | 33.3% (19) | 52.5% (31) |
| Unknown/prefer not to answer | 18.1% (21) | 17.5% (10) | 18.6% (11) |
| *Relationship status* |  |  |  |
| Partnered, living together | 31.9% (37) | 42.1% (24) | 22.0% (13) |
| Partnered, not living together | 13.8% (16) | 17.5% (10) | 10.2% (6) |
| Single | 42.2% (49) | 24.6% (14) | 59.3% (35) |
| Other/unknown/prefer not to answer | 12.1% (14) | 15.8% (9) | 8.5% (5) |
| *Everyday discrimination experience* |  |  |  |
| Never | 17.2% (20) | 19.3% (11) | 15.3% (9) |
| Rarely | 25.9% (30) | 28.1% (16) | 23.7% (14) |
| Sometimes | 36.2% (42) | 35.1% (20) | 37.3% (22) |
| Often | 16.4% (19) | 15.8% (9) | 16.9% (10) |
| Unknown/prefer not to answer | 4.3% (5) | 1.8% (1) | 6.8% (4) |
| *Prenatal care discrimination experience* |  |  |  |
| Never | 42.2% (49) | 31.6% (18) | 52.5% (31) |
| Rarely | 19.0% (22) | 26.3% (15) | 11.9% (7) |
| Sometimes | 22.4% (26) | 35.1% (20) | 10.2% (6) |
| Often | 6.9% (8) | 5.3% (3) | 8.5% (5) |
| Unknown/prefer not to answer | 9.5% (11) | 1.8% (1) | 16.9% (10) |

### Quantitative Analysis

The analytic sample includes 116 participants, excluding four participants deemed ineligible after reviewing their demographic data. We first examined trust scores and the covariates using descriptive statistics (means for continuous variables and percentages for categorical variables), followed by bivariate associations between trust scores and the covariates using ordinary least squares (OLS) linear regressions, adjusting standard errors to account for clustering by the number of events attended. Variables with p-values less than 0.05 were then included in the final multivariate model, which was refined by assessing collinearity and model fit. To account for clustering due to some participants providing multiple responses, we estimated linear mixed effects models. We conducted sensitivity analyses by excluding missing data and duplicate responses (subsequent survey responses from the same participant). All analyses were conducted in STATA (version 14) (52).

### Qualitative Analysis

We employed a thematic analysis approach (53). The qualitative lead (OJO) developed an initial deductive codebook based on the interview guide. Six analysts (OJO, PD, KV, EK, HS, and KM) then independently coded the transcripts using Dedoose software (54). Inter-rater reliability was ensured through collaborative coding of two transcripts, refining the codebook, and incorporating inductive codes. The remaining transcripts were then coded by balanced pairs of analysts, with the qualitative lead reviewing the coding for consistency. Codes related to trust were synthesized into a thematic table with five categories: 1) Trust in the health system; 2) distrust in the health system; 3) trust in CBOs; 4) distrust in CBOs; and 5) perceptions of the health system’s involvement in PV.

## RESULTS

### Quantitative results

#### Participant Characteristics

The sociodemographic and obstetric health characteristics of the participants are detailed in Table 1. About half of the participants (49%) indicated that they were currently or recently pregnant, and their age ranged from 17 to 76: 27% were aged 25 - 34, 22% were 35-44, and 26% were 45 or older. Forty-one percent identified as Black and 36% as Latine. Also, 42% reported being single, and 32% lived with a romantic partner. Educational attainment varied, with 18% not finishing high school and 45% having post-secondary education. About two-thirds reported unemployment (64%), with 47% receiving income assistance, and about two-thirds had public health insurance (66%). About one in five had experienced at least one preterm birth (20%), and one in four had experienced a previous pregnancy loss (25%).

### Trust in PV and associated factors

The sample’s standardized mean trust in PV score was 85.9 (SD = 18.9) overall, 83.5 (SD = 18.5) for pregnant and postpartum recipients, and 88.4 (SD = 19.1) among family members, (p = 0.170) (see Table 2). The mean trust score for Black participants was 91.5 (SD = 13.4), compared to a mean trust score of 81.9 (SD = 21.2) for participants from other racial and ethnic groups, (p = 0.0071).

**Table 2.** Distribution of standardized scale scores for Trust in PV.

|  | *N* | *Score* | *SD* | *Minimum Score* | *Maximum Score* |
| --- | --- | --- | --- | --- | --- |
| Combined Sample | 113 | 85.9 | 18.9 | 0.0 | 100.0 |
| Pregnant/postpartum | 57 | 83.5 | 18.5 | 0.0 | 100.0 |
| Family member | 56 | 88.4 | 19.1 | 0.0 | 100.0 |
| Black participants | 47 | 91.5 | 13.4 | 66.7 | 100.0 |
| Participants from all other racial/ethnic groups | 66 | 81.9 | 21.2 | 0.0 | 100.0 |

Abbreviations: SD: standard deviation

Table 3 shows the bivariate results for trust scores from the main sample, with subgroup analyses (pregnant/postpartum individuals and family members) in Additional file 2. Compared to Black individuals, English speakers, and those proficient in English, Latine individuals (95% CI: -21.6 - -2.9), Spanish speakers (95% CI: -21.9 - -6.8), and those with limited English proficiency (95% CI: -27.7- -1.1), respectively, had significantly lower trust in PV. Also, participants aged 25 to 44, with at least three births, one preterm birth, and those who occasionally experienced food insecurity had lower trust in PV compared to those aged 15 to 24, no prior birth, no preterm birth history, and no food insecurity, respectively. Conversely, higher trust was observed among participants who did not disclose their gender, had some higher education, and did not disclose their social support status, compared to those identifying as female, high school graduates, and with social support, respectively. There were no statistically significant predictors in the final multivariate model (Table 4).

**Table 3.**
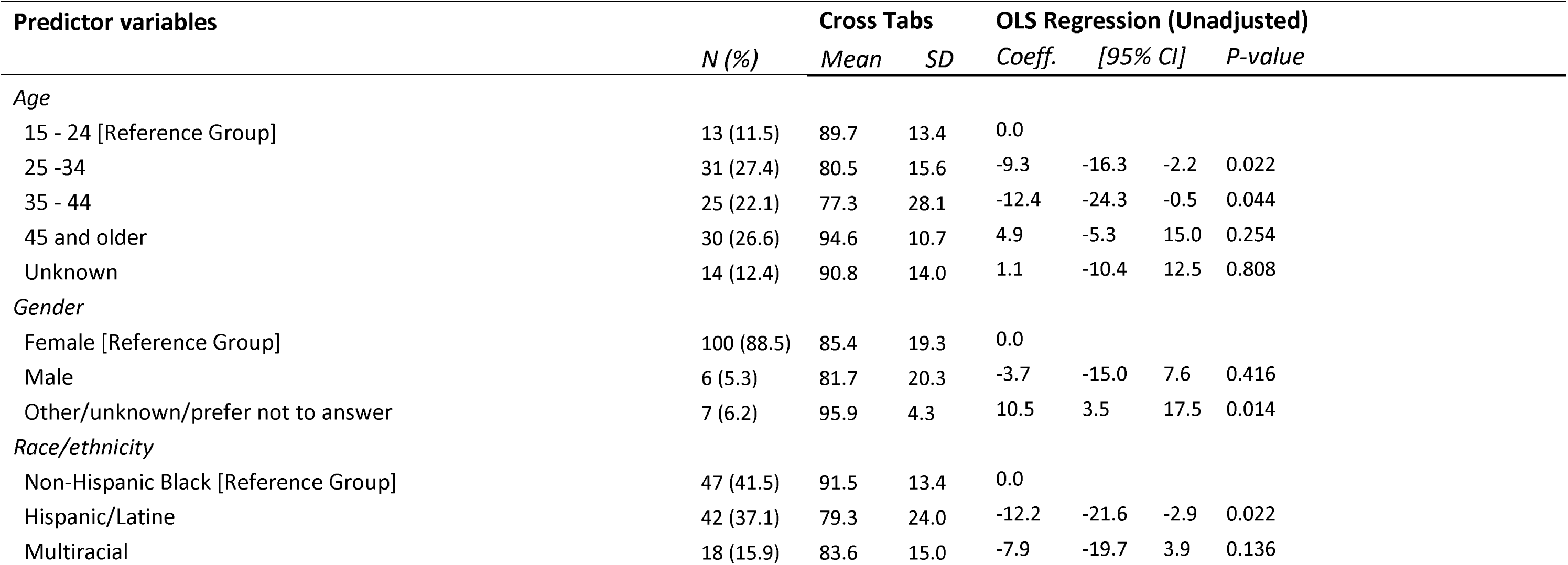
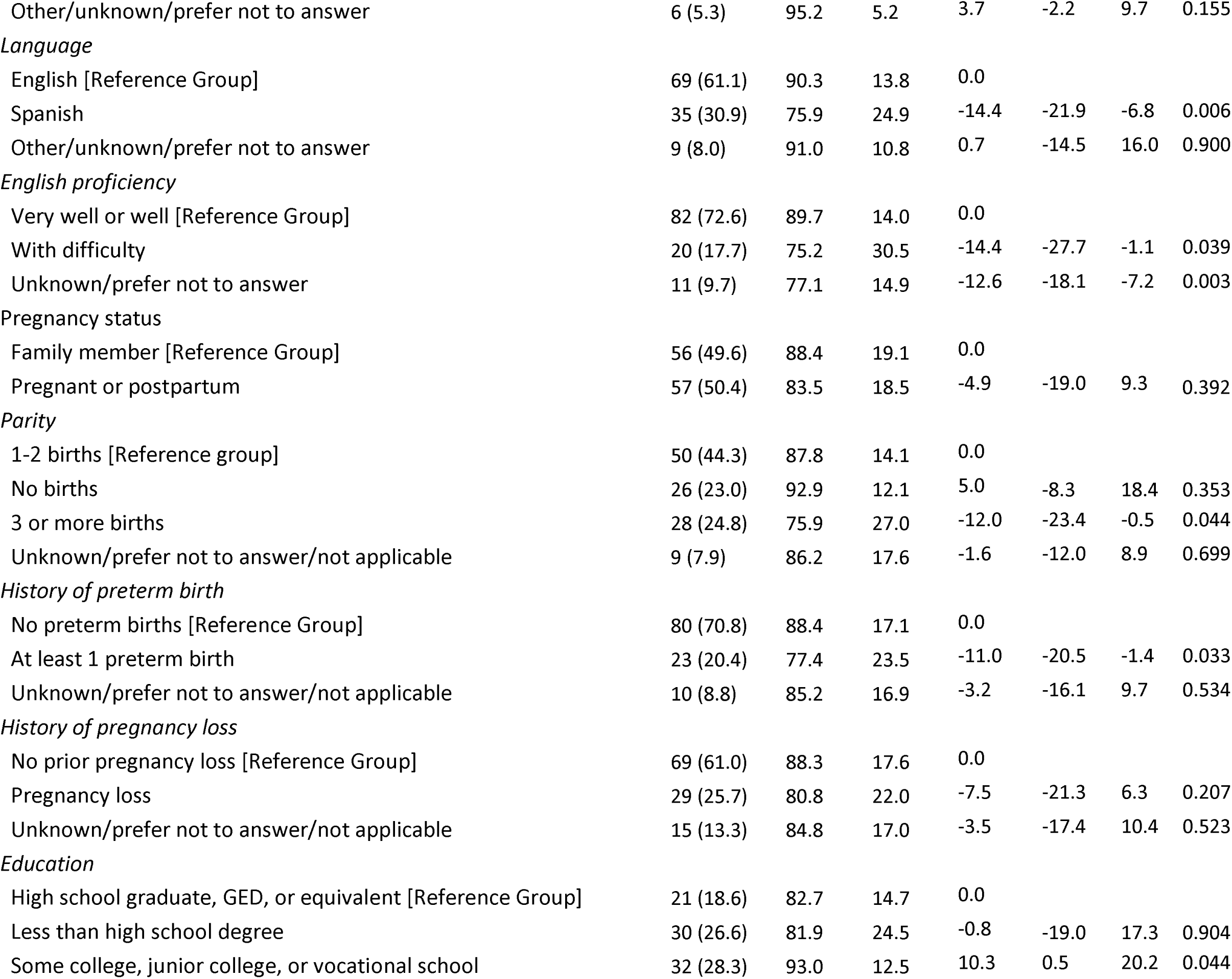
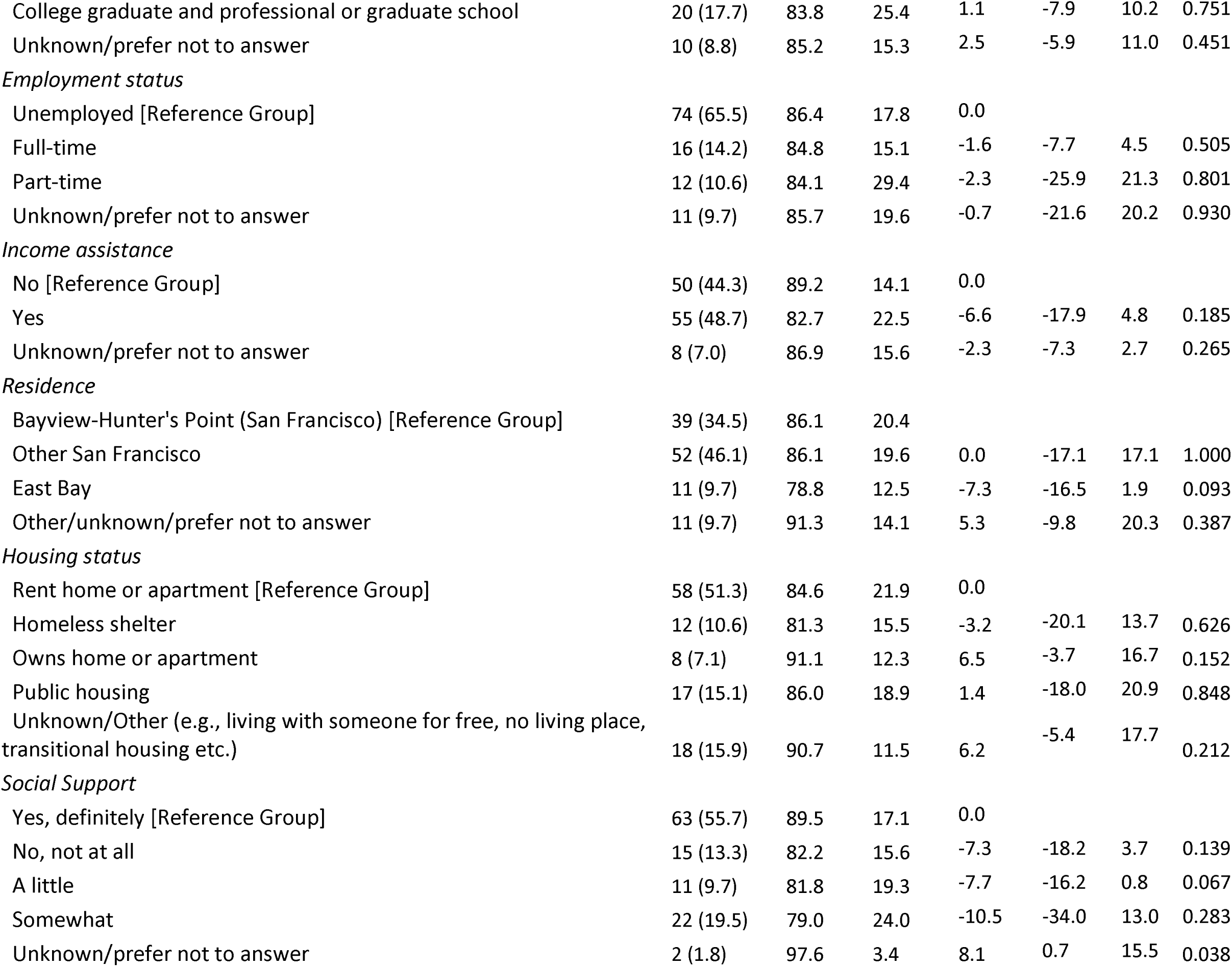
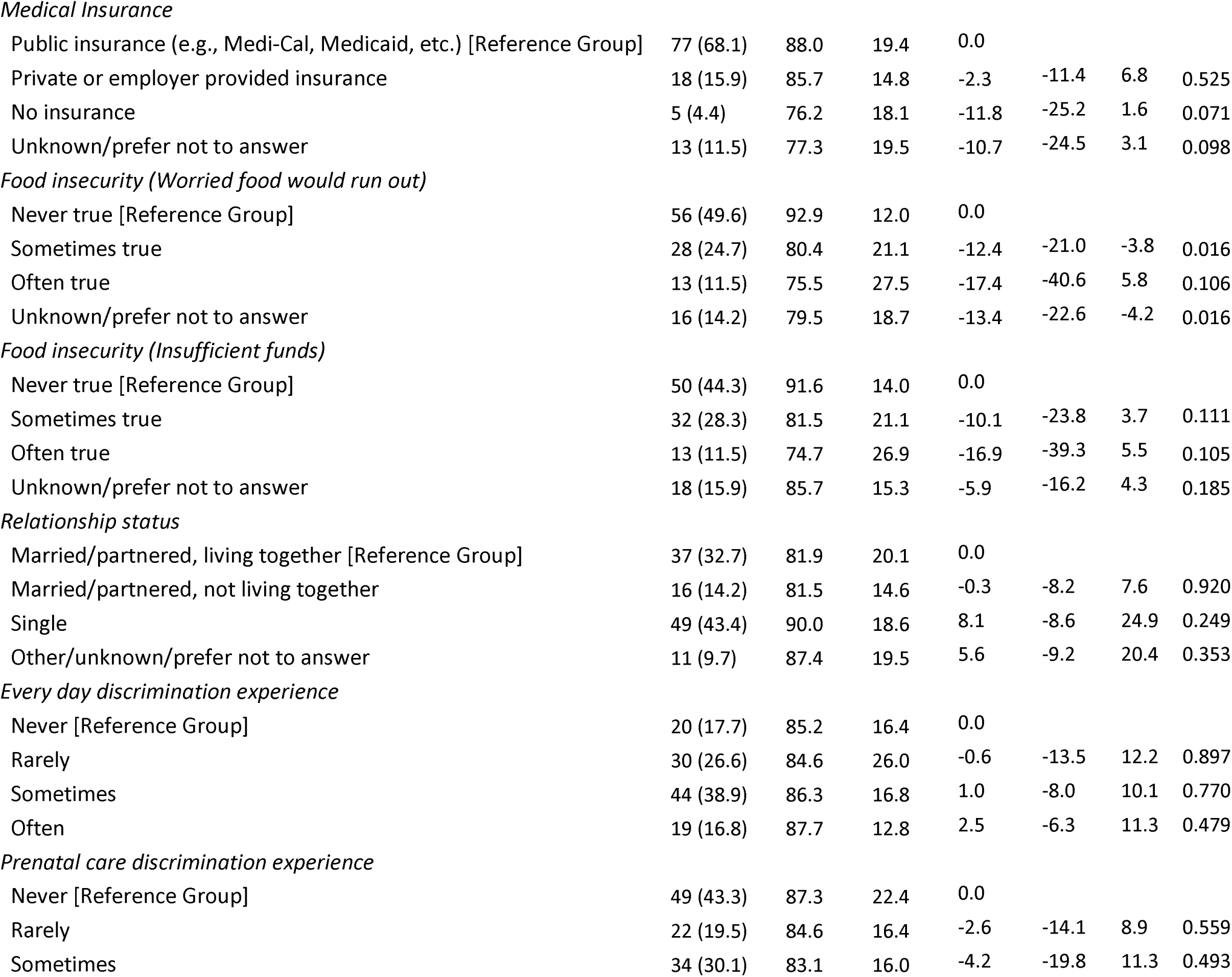
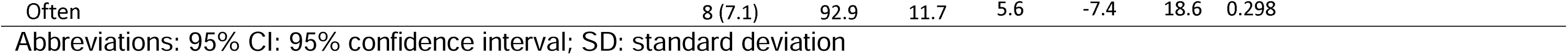
Bivariate results for predictors of Trust in PV scores.

**Table 4.** Mixed-effects multiple linear regression analysis of select predictors on trust in PV scores.

| Predictor variables | Coeff. | [95% CI] |  | P-value |
| --- | --- | --- | --- | --- |
| Age |  |  |  |  |
| 15 - 24 [Reference Group] | 0.0 |  |  |  |
| 25 -34 | -7.1 | -19.0 | 4.8 | 0.240 |
| 35 - 44 | -5.1 | -17.0 | 6.8 | 0.402 |
| 45 and older | 1.6 | -10.6 | 13.8 | 0.795 |
| Unknown | -0.1 | -13.5 | 13.3 | 0.986 |
| Gender |  |  |  |  |
| Female [Reference Group] | 0.0 |  |  |  |
| Male | 4.3 | -23.6 | 32.1 | 0.764 |
| Other/unknown/prefer not to answer | 6.8 | -10.0 | 23.5 | 0.427 |
| Race/ethnicity |  |  |  |  |
| Non-Hispanic Black [Reference Group] | 0.0 |  |  |  |
| Hispanic/Latine | 13.0 | -2.9 | 28.9 | 0.109 |
| Multiracial | 0.3 | -9.4 | 10.1 | 0.945 |
| Other/unknown/prefer not to answer | 8.2 | -10.4 | 26.8 | 0.390 |
| Language |  |  |  |  |
| English [Reference Group] | 0.0 |  |  |  |
| Spanish | -11.5 | -29.9 | 6.8 | 0.218 |
| Other/unknown/prefer not to answer | -1.3 | -16.8 | 14.3 | 0.871 |
| English proficiency |  |  |  |  |
| Very well or well [Reference Group] | 0.0 |  |  |  |
| With difficulty | -9.0 | -22.8 | 4.8 | 0.203 |
| Unknown/prefer not to answer | -11.0 | -26.6 | 4.5 | 0.163 |
| *History of preterm birth* |  |  |  |  |
| No preterm births [Reference Group] | 0.0 |  |  |  |
| At least 1 preterm birth | -6.8 | -14.9 | 1.2 | 0.096 |
| Unknown/prefer not to answer/not applicable | -2.1 | -27.3 | 23.2 | 0.873 |
| *Education* |  |  |  |  |
| High school graduate, GED, or equivalent [Reference Group] | 0.0 |  |  |  |
| Less than high school degree | 4.5 | -5.5 | 14.4 | 0.380 |
| Some college, junior college, or vocational school | 4.4 | -4.9 | 13.6 | 0.355 |
| College graduate and professional or graduate school | -4.6 | -14.7 | 5.6 | 0.379 |
| Unknown/prefer not to answer | 1.0 | -11.8 | 13.9 | 0.875 |
| *Food insecurity (Worried food would run out)* |  |  |  |  |
| Never true [Reference Group] | 0.0 |  |  |  |
| Sometimes true | -7.2 | -15.9 | 1.5 | 0.104 |
| Often true | -11.1 | -22.7 | 0.4 | 0.058 |
| Unknown/prefer not to answer | -8.5 | -19.5 | 2.5 | 0.128 |
| *Social Support* |  |  |  |  |
| Yes, definitely [Reference Group] | 0.0 |  |  |  |
| No, not at all | -2.3 | -12.8 | 8.1 | 0.658 |
| A little | -6.4 | -19.5 | 6.7 | 0.340 |
| Somewhat | -6.0 | -14.0 | 2.0 | 0.144 |
| Unknown/prefer not to answer | 9.3 | -33.4 | 52.0 | 0.669 |
| *Mean dependent var* | 85.883 |  |  |  |
| *Number of observations* | 113 |  |  |  |
| *Prob > chi2* | 0.000 |  |  |  |
| *SD dependent var* | 18.876 |  |  |  |
| *Chi-square* | 61.001 |  |  |  |
| *Akaike crit. (AIC)* | 992.86 |  |  |  |

Abbreviations: 95% CI: 95% confidence interval; SD: standard deviation

### Sensitivity analyses

A sensitivity analysis, which excluded missing responses and limited participant engagement to their first PV encounter, yielded nearly identical standardized mean trust scores: 83.4 (SD = 19.8) for the main sample (N = 88), 82.7 (SD = 19.0) for pregnant and postpartum recipients (n = 47), and 84.3 (SD = 20.9) for family members (n = 41).

### Qualitative results

Qualitative findings are organized around three key areas: factors influencing trust in the health system, factors shaping trust in CBOs, and perceptions of health system involvement in PV. In general, participants expressed mixed levels of trust in the health system and high trust in CBOs. Most participants viewed the health system’s involvement in PV positively, though a few voiced skepticism about its role.

### Factors shaping trust in the health system

We identified two themes from participants’ descriptions of factors that shaped their trust in the health system: *Person-centered care and care satisfaction,* and *familiarity with the health system*.

### Person-centered care and care satisfaction

Participants attributed their trust in the health system to the positive interactions they had with health care providers, which reflected core domains of person-centered care—care that is respectful of and responsive to individuals’ preferences, needs, and values (55). This included provider responsiveness, such as provider friendliness, welcoming demeanor, and attentiveness, which reinforced trust.

> *“I think that their [X hospital] healthcare providers – I mean, their healthcare, like nurses and doctors, are really nice and welcoming. And they make sure that I know that everything’s okay. And, if anything was wrong, you know, they would let me know. And they’re just really welcoming and nice.”* —Multiracial pregnant participant, less than 30 years old

Trust was also tied to effective communication with participants, noting confidence in provider knowledge, comfort asking questions, responsiveness to their questions, and positive reassurances:

> *“I trust them* [health system] *just because personally, the doctor I work with, with my daughter, I feel very confident with her knowledge, and I am comfortable asking her any questions. I’m never hesitant to ask, and she’s always there to respond and kind of reassure me if I have any concerns or anything about my daughter. But I guess it’s just more the knowledge that she has that I’m very confident about.” —*Latine postpartum participant, less than 30 years old
>
> *“Maybe because, what they* [healthcare providers] *teach me, it turns out to be true. …So, whatever they teach me, they tell me this is going to happen, this might happen, do you feel this, or you’re going to feel this after a month, or you’re going to feel this when the baby comes – that stuff. Everything they teach me turns out to be true. And they are always positive. Like, even if I am in pain, “Oh, don’t worry. You going to be fine. …. No. I am not fine. Especially at the delivery time. [laughs] It’s hard. …. Yeah. But you’re going to be fine after this – just hours after this pain. And then, you know when I forget the pain, I was like, “Wow. They were right.” [laughs] Yeah. It’s only over for the delivery.” —*Black Pregnant participant, between 30 and 44 years old

In addition, autonomy in decision-making was tied to trust, with some participants noting that their trust in the health system stemmed from being empowered to make choices regarding their health, including changing physicians with whom they are dissatisfied:

> *“Because they* [health institution] *give you the opportunity to say “hey, I don’t like this, I don’t think this provider, meaning doctor, is showing my best interest, or* [acknowledging] *my thoughts on the way I want my healthcare to be.” They give you the right to change doctors, to change providers. So, I have to trust, you know, that this institution has given me the right to choose how my health is going to be. Because if they hadn’t given me the right to choose, I probably wouldn’t be pregnant now because my first OB/GYN was telling me that I should have weight loss surgery before I tried to have another child […] I told her* [new provider] *my concerns, and you know, and after a couple weeks of me being with her, I would never tell my first OB/GYN when I got pregnant again; I just immediately found myself another one, because I didn’t want to hear the negative thoughts that she was giving, and I didn’t want me to be stressed. With this new doctor, I haven’t been [stressed]. She listens to me, she calls me, her intern calls me and checks on me.” —*Black pregnant participant, between 30 and 44 years old

Further, participants’ trust in the health system was linked to their perception of healthcare providers being in tune with their unique circumstances and making decisions based on that, in a way that increased their satisfaction with services received. For instance, one participant valued the exception made during her childbirth, allowing her to stay in the hospital despite not being adequately dilated at presentation. She appreciated this decision as it spared her from going home, where she lacked social support and feared suffering alone:

> *“I was nervous [of giving birth] because I didn’t know what to expect, and I was by myself initially […]. When I first went to the hospital, I wasn’t even dilated yet, and they were going to send me home. But when I was at home, I was having really bad contractions. And so, they made an exception. They’re like, “We’ll go ahead and keep you.” And I know they didn’t have to do that. Because I was zero centimeters dilated, even though I was having crazy contractions. And I know people that they’ve sent home that are three centimeters dilated, or four centimeters dilated, and they send you home. But for whatever reason, they had grace on me, and they let me stay. Because it was really hard to be here going through labor in my room. It was very intense. And I wanted to be close to the medical attention. And I didn’t want to go home and have to come right back. So, I was thankful that they made the exception for me, because I didn’t want to come home and suffer all by myself. It was really bad. But, I made it! And I really like their facility. I had a view of the ocean and the ships on the ocean.” —*Latine postpartum participant, between 30 and 44 years old

### Familiarity with the health system

One participant attributed their trust in the health system to long-term engagement, highlighting the system’s consistent presence in her life:

> *“I was born there* [X hospital]. *I’ve been there on and off for all my life. So, I really don’t know no other hospitals but X hospital […] I’ve been dealing with them, like I said, all my life. That’s the hospital I know.”* —Black family member, 45 years old and above

### Factors shaping distrust in the health system

We identified three main themes from participants’ reports of factors shaping their distrust in the health system: *Experiences of racism and discrimination*, *Health system unwilling to center Black women’s health, and Lack of preventative care*.

### Experiences of racism and discrimination

A few participants expressed distrust of the health system, citing personal or familial experiences of racism and discrimination, as well as the systemic racism faced by the Black community when accessing its services:

> *“Because I know when I was born – like my siblings, most of us were born in X Hospital and my mom did experience a lot of racism, again that was the 80s, so hopefully I’m thinking things have changed. There is a caution, and I don’t want to speak for the whole Black community, but I know that I have and a lot of people I know who are Black, especially women when it comes to like doctors and our trust to the system, hence why we don’t see people getting vaccinated.”* —Black pregnant participant, between 30 and 44 years old

### Health system unwilling to center Black women’s health

A few participants expressed distrust in the health system, noting that it often neglects the health needs of Black women and fails to center them in the development of treatment plans. For instance, one participant highlighted the lack of adequate research on Black women’s health, which limits the system’s ability to effectively support their well-being:

> *No. I don’t trust [laughs] hospitals, especially as a Black woman. I don’t trust them. They’re always telling me something about myself. You know, you go to get care for one thing, and then, they’re like, “Oh. Well, first of all, you need to lose weight.” Okay. It’s like, okay, beyond that, what can you do to help me? Like don’t center my weight as like the main focus of your health intervention. Because I just don’t feel like they have enough research being done that centers Black women to make me feel comfortable that they’re going to be centering me in the medical office. There’s just not enough research centered on Black women and their physical bodies and experiences and things like that. So, until that happens, I’m just not – [laughs] I don’t trust – I mean I’ll use it, but I don’t trust it.” —*Black postpartum participant, 45 years old and above

### Lack of preventative care

Participants criticized the system’s focus on crisis intervention rather than prevention, arguing that it is primarily designed to act only during crises or after conditions have worsened:

> *“Because I don’t think the medical system is set up to help us prevent, but more so to intervene in a crisis. From my first pregnancy, the experience that I had with going to check-ups with [X hospital] and my homebirth team – the [X hospital] side was so negative because they would constantly talk about things that could happen and bad things, and the focus was so much on instilling fear.” —*Pregnant participant, less than 30 years old, race/ethnicity unknown

### Factors shaping trust in Community-based organizations

Most participants expressed a strong trust in CBOs to support their health and well-being, with some indicating greater trust in CBOs than in the health system. This was attributed to several factors, including CBOs’ provision of *person-centered care*, their *holistic care approach*, their *relatability*, and their *commitment to meet community’s needs*. Additionally, participants’ long-standing relationships with these organizations were key in fostering trust.

### Provision of person-centered care

Participants highlighted that the nature of care they received from CBOs was a key factor in building their trust in these organizations. Their descriptions reflected several domains of person-centered care marked by responsive and compassionate staff who served as dedicated supporters and advocates. For example, one Black postpartum participant described how her doula played a pivotal role, offering unwavering support throughout her pregnancy and passionately advocating for her during her hospital-based childbirth. This advocacy empowered the participant and facilitated a trauma-free birth experience by ensuring that the care provided aligned with the participant’s birthing preferences:

> *“Because I did end up working with a doula. …. That, I think, was my saving grace because she just was like, “Girl, you don’t need to be induced.” […] “You got pregnant naturally […], “That means your body was healthy and ready to have a baby. So, you can have this baby naturally” […] I got railroaded at the very end of my pregnancy by my healthcare providers trying to get me to agree to an induction. At one point, a nurse and my OB had double-teamed me while I was on my non-stress test, telling me about all of the risks of not getting induced. I [would] have agreed to be induced, 100 percent, had I not had the doula. It would have been the difference between me having a completely natural childbirth versus having a traumatic childbirth […] Instead, I worked with the doula. She had me laboring at home with my partner for most of the time […] When I got to the hospital, I was nine-and-a-half centimeters dilated. I was so far into my pregnancy [close to birth] that they couldn’t do any interventions. All they could do was barely put me on the labor and delivery table. And he came out in two pushes.” —*Black postpartum participant, 45 years old and above

Participants also noted that CBOs excelled in clear, informed communication with individuals, families, and the community. They emphasized the organization’s ability to be attentive, actively listen, and respond to the needs of the community.

> *“I would trust them almost a hundred percent more than the hospital healthcare system. Because they’re community-based and they are trained and attuned to listening to the community members. That’s the difference. It’s about how you listen. And the medical community – I mean, for all intents and purposes – has a God complex. They think they know everything. And they’re not great listeners.” —*Black postpartum participant, 45 years old and above

One participant expressed appreciation for the respect shown by CBOs, particularly valuing the confidentiality of their discussions. They considered this trustworthiness a crucial aspect of their relationship with these organizations:

> *“I think they* [CBOs*] are trusting. Because I feel like, whenever I ask them* [CBOs] *something or I tell them something, they really just keep it confidential, and it’s always just between me and them. I never feel like there’s someone else in the conversation.” —* Multiracial pregnant participant, less than 30 years old

### Holistic approach to care

Participants highlighted that CBOs earned their trust by prioritizing a holistic approach to care. These organizations focused on overall health and wellness, rather than relying on extensive medical interventions. Unlike the health system, CBOs were not constrained in their ability to provide comprehensive, personalized care:

> “*They are not bound to the standards or rules that medical providers are necessarily bound to by the medical system. So, they can look at more holistic approaches and – yes, less medical intervention, but more medical prevention*.” *—*Pregnant participant, less than 30 years old, race/ethnicity unknown
>
> *“I mean, I’ve been going out there to X CBO when I need acupuncture. If I need something, they tell me what to do, you know, what kind of treatment to use, and all this stuff. Like the needles and all this stuff. Like my shoulder, I can’t lift my shoulder. Yeah, they tell me what treatment to use*.” *—*Black family member, 45 years old and above

### Relatability and approachability

Participants noted that CBOs were more relatable and approachable than the health system. Some compared their interaction with CBOs to a familial bond, which fostered a sense of comfort and encouraged open communication and problem-sharing:

> *“Because, you know, for example, the hospital – X Hospital is like you are dealing with the doctors, nurses, and professionals. But, when we come to the communities* [CBOs], *I feel like, when we come to the communities* [CBOs], *you’re dealing with a mother like you […] And, with different perspective people, it’s even easier to approach. You know? Instead of dealing with upper – I feel like – let’s call them “upper class.” But this is like siblings, like sisters. […] So it feels easier to talk to them, you know, to share your problems,” —*Black Pregnant participant, between 30 and 44 years old

### Reliability and commitment

Participants expressed trust in CBOs for their reliability and consistent availability in meeting their needs:

> *“I trust them because they’re always able to provide the help that I need. I guess with the two ones that I rely on, which are the material stuff - just like the diapers and wipes and the formula and my baby’s food.” —*Latine postpartum participant, less than 30 years old

They also valued the organizations’ deep commitment to the community, noting how CBOs go above and beyond to meet individuals where they are and provide tailored support.

> *“At this point, I do trust them. Because they* [CBOs] *could be doing something else with their time on a Saturday. They could be elsewhere with their family. But obviously, this might be something that they believe in, and they have a heart for it. And to give up your time, wow, that’s a lot. Time is something you can’t ever get back. You can’t get that back. So, that’s very special for me that they would donate their time like that.” —*Latine postpartum participant, between 30 and 44 years old

### Familiarity

Participants attributed their trust in CBOs to their familiarity with the organizations and the long-standing relationships they had built. This trust was further reinforced by their positive experiences with these organizations:

> *“I’ve been going there for a long time, and they’ve been known to me, so I trust them.” —*Black family member, 45 years old and above
>
> *“I think so [I trust them]. Those [CBOs] are the places that I know. Because certainly, in the past, they helped me a little bit. They had the resources that I need.” —*Latine pregnant participant, between 30 and 44 years old

### Skepticism of CBOs

While no participant expressed complete mistrust of CBOs, some mentioned reasons for their skepticism. These included concerns about CBOs’ unresponsiveness to community needs and an insufficient number of CBOs to meet growing community demand:

> *“I don’t know* [if I can trust CBOs] *because I’m still waiting on a follow-up. So, I don’t know. I don’t know if I can trust them for anything, not even a callback. So, I don’t know.” —*Black pregnant participant, less than 30 years old
>
> *“And so, there’s not enough community-based agencies here. They’re trying to get some together. There’s a medical building on Third Street. There’s dental – there’s a X community clinic that’s being rebuilt and all. So they’re progressing. But they’re nowhere near what could be, and people don’t have to go across town to get services if they can get them in this community.” —*Black family member, 45 years old and older

Also, one participant stated that they didn’t fully trust CBOs because they had a general lack of familiarity and understanding of the organizations:

> *“I really don’t know too much about them, so I don’t think I’ll trust them as much. I don’t know nothing really about them. So, I can’t trust them. If I don’t know nothing about you, I can’t put my trust in you.” —*Black family member, 45 years old and older

### Perceptions of the health system’s involvement in PV

Given the extensive corpus of literature on mistrust in the health system, we asked participants about their perceptions of the health system’s involvement in PV to help understand trust in PV. Overall, participants had a positive outlook on the health system’s involvement in PV. This was based on *prior positive experiences with the health system, the health system’s role in providing resources and information*, and *meeting community needs*.

### Positive experiences with the health system

Participants attributed their positive perceptions of the health system’s involvement in PV to their own positive experiences with the system:

> *“I feel good about it* [health system’s involvement in PV]*. I mean, it’s part of wellness. And I gave birth at X hospital, and I had a great experience there, too. […] I feel good about it, definitely.” —*Latine postpartum participant, 30-44 years old
>
> *“I feel confident just because I did have services happen in X hospital. I only had one, but from my experience with that, it went very well.” —*Latine Postpartum, less than 30 years old

### Health system’s role in providing resources and information

Participants valued the health system’s involvement in PV, recognizing its role in providing vital information and resources:

> “*It* [health system’s involvement in PV] *feels good, because they are two very important and large institutions, and because they have information and resources that they can provide to us.” —*Latine postpartum participant, between 30 and 44 years old
>
> *“Because they* [the health system] *have a different perspective, as well as knowledge. Both hospitals have different knowledge, and they can bring different ideas and different thoughts to the table. So, it’s good to have different resources and different people working together because everybody’s got different opinions and different ideas. So, it’s good that they both joined to help with the services of the Pop-Up.” —*Black pregnant participant, less than 30 years old

A participant further noted that the health system’s involvement in PV acts as a critical resource-information bridge for the community, with hospitals and clinics serving as key sources of knowledge about sociomedical interventions like PV. The participant shared that she only learned about a CBO that supports Black pregnant people*—*and serves as a PV partnering organization*—*through one of her healthcare providers:

> *“I feel good. I’m surprised a lot more major health providers aren’t a part of it. Because there are pregnant people that go to X hospital, even though it is a private company. But still, I’m surprised that you know, they’re, they’re not a part of it. I’m surprised X hospital is not a part of it […] I’m just really surprised that a lot more major hospitals aren’t a part of it. I would* [like more hospitals to be involved], *because that’s how you get the information out there. You know, that’s how you spread the word, that’s how you are more involved in your community, because had it not been for the intern that was my therapist at X hospital, I would have never known about Black Infant Health.” —*Black pregnant participant, between 30 and 44 years old

### Meeting community needs

Some participants supported the health system’s involvement in PV, noting the high demand for clinical services in their community and advocating for greater health system engagement to better meet these needs:

> *“That’s good. We need more hospitals involved. We’ve got sick children. We’ve got lupus. We need all of our hospitals to be involved. Not just two or three. All of them. That’s how I feel… people don’t just go to X and X hospital. They go to all these hospitals around San Francisco. You don’t go to just two. People go to different hospitals. I think all the hospitals should be involved and let their patients know.” —* Black family member, 45 years old and older

One participant noted that the health system’s involvement in PV demonstrates its recognition of the community’s need for clinical services and reflects its support for the community:

> *“I think it’s pretty awesome that they’re coming together and noticing that these are the things that women need… Why is it important for them to be involved? To let us know that there are, you know, health providers or doctors that are on our side and notice that this is something that we struggle majority of the time while being pregnant or after being pregnant. So having these services done by them, I guess, is a plus and a must.” —* Multiracial pregnant participant, between 30 and 44 years old

### Reservations about the health system’s involvement in PV

While most participants appreciated the health system’s involvement in PV, a few expressed concerns, emphasizing the importance of the system acknowledging and addressing its role in systemic issues. They also emphasized the importance of the health system understanding its position within the broader context of mistrust in the Black community.

> “*I actually think that that is fine and good, as long as they recognize that they are part of the problem* [laughs] *and are working towards like becoming a part of a solution.” —* Black postpartum participant, 45 years old and above
>
> *“That’s the fact that they* [the health system] *are involved shows […] There is a caution, and I don’t want to speak for the whole Black community, but I know that I have and a lot of people I know who are Black, especially women when it comes to doctors and our trust to the system […]There’s a mistrust in this country, and I think it’s based and it’s valid […] when it comes to the medical field and how they have interacted with Black people in this country*.” *—*Black pregnant participant, between 30 and 44 years old

## DISCUSSION

We aimed to assess individuals’ perceptions of trust in PV and explore their trust perceptions of the health system and CBOs, as well as the health system’s involvement in PV. Trust in the PV model was generally high among participants and was influenced by factors such as race and ethnicity, language, history of preterm birth, and food insecurity, as found in the bivariate analyses. Qualitative findings revealed that trust in both the health system and CBOs was shaped by experiences of person-centered care and organizational familiarity. Trust in CBOs was additionally influenced by their emphasis on holistic care, relatability, and responsiveness to community needs. Conversely, distrust in the health system stemmed from experiences of racism and discrimination, inadequate preventative care, and the system’s neglect of Black women’s health. Participants held mixed views on the health system’s role in PV, with most highlighting its positive contributions, while a few voiced skepticism due to ongoing structural racism and inequities in care.

This study is the first to use the trust scale to assess trust in care from a community and institutional partnership. Although not directly comparable given the adaptations we made, the trust scores for PV are generally higher than scores obtained using the trust scale in healthcare settings (51). The high levels of trust in PV are likely linked to PV’s cross-sector collaborative approach to care delivery that includes known trusted sources such as CBOs. Research also shows that such cross-sector partnerships promote knowledge sharing and support the development, integration, and dissemination of diverse information, resources, and care delivery approaches (56). These collaborations often result in services that are more relevant, culturally affirming, and responsive to community needs (57), thereby fostering greater trust in the model.

Although the associations between race and ethnicity, language, history of preterm birth, food insecurity, and trust did not remain significant in the multivariate analysis, they are meaningful and supported by qualitative data. First, because PV centers Black families, services are focused on Black families’ experiences, and initial challenges with translators affected the ability of non-English speaking participants to effectively navigate the Village (47). This may have contributed to higher mistrust among Latine and Spanish-speaking individuals. These findings highlight the critical importance of addressing language barriers when assessing racial and ethnic disparities in PV.

Second, a history of preterm birth is a traumatic experience, due to the risk of severe infant injury or death (58), and has potentially profound and lasting effects on family well-being. Nearly half of mothers and a third of fathers experience post-traumatic stress disorder (PTSD) following a preterm birth (59). For women of color, this is often compounded by negative healthcare experiences, including disrespectful care and unmet informational needs (60). The cumulative impact of this trauma—of the preterm birth itself and systemic mistreatment—can deepen distrust toward the health system. While PV aims to provide person-centered care, lower trust among those with a history of preterm birth may reflect inadequate delivery of trauma-informed care at PV. Therefore, training providers to recognize and address trauma-related needs is crucial to prevent unintentional re-traumatization (61).

Additionally, the association between food insecurity and trust may be understood through the lens of Maslow’s hierarchy of needs: individuals must first satisfy basic physiological and safety needs—such as food and housing—before they can pursue higher-order needs such as belonging or self-actualization (62). Thus, when these basic needs are unmet, attention and resources are often directed toward survival needs at the expense of being able or willing to engage with services perceived as peripheral to their unmet urgent needs. Although food was provided at PV events, this may not have been sufficient to meet the needs of individuals experiencing food insecurity. Consequently, if PV is not perceived as addressing these immediate needs, they may be less inclined to trust the model.

That trust in the health system and CBOs is linked to person-centered care and service satisfaction is unsurprising. Transparent communication fosters shared decision-making, empowers individuals, and builds trust (63,64). Additionally, patient satisfaction—driven by provider responsiveness, clear explanations, and respectful interactions—is a key contributor to trust (65,66). Prior research shows that staff attitudes are the strongest predictor of satisfaction (67), and nearly half of Black pregnant women’s satisfaction with prenatal care is attributed to provider communication, including explanations, empowerment, and interpersonal style, such as friendliness and perceived discrimination (68). The importance of person-centered care—characterized by respectful, open communication and shared decision-making—in fostering trust is therefore evident.

Trust in CBOs, unlike the health system, was also largely attributed to CBOs’ prioritization of holistic care, relatability, and commitment to addressing community needs. CBOs are often deeply rooted in the communities they serve, giving them a nuanced understanding of how social determinants of health—such as race and ethnicity, education, and income—interact to impact health and well-being (69). This allows them to deliver services that extend beyond clinical care and address structural inequities (70,71). Further, their holistic, personalized approach, grounded in lived experience, may foster trust. CBOs also often benefit from having staff and organizational leaders who share similar cultural and socioeconomic backgrounds with the communities they serve, creating mutual understanding and comfort; this relatability may enhance feelings of respect, belonging, safety, and trust (72). Moreover, CBOs often serve as advocates for minoritized communities, advocating for resources, policy reforms, and social justice (73). As a result, minoritized individuals may be more inclined to trust organizations that share their values, actively strive to meet their needs, and address structural inequities (35,74).

The association between experiences of racism and discrimination and lower trust in the health system is well-documented in the literature (75). This distrust stems from the complex interplay of individual, interpersonal, institutional, and structural racism. At the individual level, racist beliefs and ideologies can manifest as interpersonal racism, leading healthcare providers, influenced by implicit biases, to treat Black patients poorly, such as perceiving them as less compliant with medical directives (76) and dismissing their pain and other concerns (77). These ingrained racist ideologies and practices are further reinforced at the institutional level, leading to significant disparities in care quality (60,78,79). Ultimately, interpersonal and institutional racism converge to form structural racism, where racist ideas and beliefs are enshrined in laws and policies that allocate resources in ways that disempower and dehumanize historically minoritized populations (80,81), perpetuating unequal access to high-quality care and poorer maternal and infant health outcomes. These all contribute to distrust of the healthcare system.

While recent initiatives, such as implicit bias training, have aimed to reduce discrimination and build trust in the health system, they are insufficient alone. Addressing racial health inequities requires bold, innovative partnerships and coordinated advocacy among health systems, community-based organizations, and government agencies (82,83). These collaborations must go beyond surface-level interventions and focus on addressing the root causes of racial health inequities—structural racism—by advocating for essential national, local, and institutional policy changes (82,83).

Given the complex history between minoritized populations and the health system, it was important to explore perceptions of the health system’s involvement in PV, as this has significant implications for building trust with these communities. Our findings revealed a duality in perspectives. Most participants viewed the health system’s involvement in PV positively, citing that it meets their needs. A possible explanation is that people are more likely to trust the health system when their health needs are met, thus a higher acceptance of the health system’s involvement in PV. However, we also found that a few individuals were skeptical of the health system’s involvement in PV, viewing it as complicit in perpetuating racial health inequities rather than a reliable partner in care. This skepticism and distrust are rooted in a long history of structural racism. The roots of this distrust can be traced back to the era of slavery, during which the suffering and exploitation of enslaved women’s bodily autonomy for the sake of science inadvertently led to the establishment of the field of gynecology (84). This legacy of medical racism continued with egregious ethical violations, such as the Havasupai Diabetes Project, where blood samples from the Havasupai Tribe were misused for unauthorized research (85), and the Tuskegee Syphilis Study, in which Black men were deliberately denied treatment to observe the natural progression of syphilis (86), leading to profound distrust in the health system. To genuinely build trust with historically minoritized populations, the health system needs to proactively recognize and confront its own biases. This requires a multi-faceted approach: openly acknowledging the historical context of medical and structural racism, providing comprehensive training for staff that is both culturally affirming and trauma-informed, actively engaging with the community and partnering with CBOs, and establishing transparent practices that show a dedication to health equity (87).

### Strengths and Limitations

This study has some limitations and strengths. First, since the evaluation took place within a real-world setting and the model’s dynamic co-creation and iteration approach enabled it to adjust to the needs expressed by people, the model’s consistency varied over time. Secondly, although our sample closely reflected the priority population, our purposive and convenience sampling methods limit the generalizability of our findings. A significant strength of this study is the adaptation of a validated quantitative trust measure, enhancing validity and reliability. Additionally, this is one of the few studies to evaluate trust in a sociomedical intervention and the first study, to our knowledge, to explore people’s perceptions of the health system’s involvement in such an intervention.

## CONCLUSIONS

The pilot implementation of the Pregnancy Village model, the SF Family & Pregnancy Pop-Up Village, fostered high levels of trust among Black and other minoritized pregnant individuals and their families in San Francisco, California. These high trust perceptions suggest that cross-sector models like PV can bridge historical divides between community members and health systems by creating spaces where healthcare institutions can interact with, learn from and collaborate with community-based organizations in real time, and can be more community-driven, responsive, and structurally accountable than may be feasible in their usual settings. The high trust perceptions highlight the importance of sustained cross-sector collaborations between community and institutions. Future research should explore longitudinal outcomes of trust-building efforts, assess scalability across diverse settings, and examine the mediating role of trust in health outcomes. Policy efforts should prioritize sustained funding and institutional-community co-leadership to ensure models like PV thrive and evolve.

## Data Availability

The datasets generated and/or analyzed during the current study are not publicly available due to privacy and ethical restrictions but are available from the corresponding author upon reasonable request.

## List of abbreviations

CBOs: community-based organizations
Pregnancy Village: PV
Post-traumatic stress disorder: PTSD

## DECLARATIONS

### Ethics approval and consent to participate

We received ethics approval from the Institutional Review Board at the University of California, San Francisco (#20–32393). Verbal informed consent was obtained at PV events after being given information about the study and before survey administration.

### Consent for publication

Not applicable.

### Authors’ contributions

OJO: Investigation, validation, methodology, data curation, formal analysis, visualization, project administration, writing – original draft, writing – review and editing; AJB and AME: Investigation, validation, methodology, data curation, project administration, writing – review and editing; CR: writing – original draft, writing – review and editing; KV: Data curation, formal analysis, writing – review and editing; KC: writing – review and editing; MAN: Conceptualization, investigation, validation, methodology, project administration, resources, funding acquisition, writing – review and editing; PAA: Conceptualization, investigation, validation, methodology, project administration, resources, funding acquisition, supervision, writing – review and editing.

## Acknowledgements

We would like to thank all PV participants, particularly those who kindly shared their experiences during the interviews. We also deeply appreciate all the organizations, staff, and volunteers who supported PV events by providing care and services to underserved residents of San Francisco. Lastly, we would like to thank all research team members, especially those who provided support in analyzing the qualitative data.

## Footnotes

1 Participants who attended several monthly PV events were permitted to fill out one survey for each event they attended. In total, 15 individuals completed the survey multiple times: one participant completed it eight times, another completed it six times, two participants completed it three times, and eleven completed it twice. Overall, 89 unique individuals participated in the evaluation.

