## Supplemental Tables for "Building trust through collaboration: A mixed-methods evaluation of San Francisco’s Pregnancy Village model of cross-sector care delivery"

**Additional file 1. Distribution of trust scale items**

|  | Total | Currently pregnant/recently pregnant | Family member |
| --- | --- | --- | --- |
|  | N=113 | n=57 | n=56 |
| 1. I believe the service providers at Pregnancy Pop-Up Village know what they are doing. |  |  |  |
| 0, Strongly disagree | 4.4% (5) | 7.0% (4) | 1.8% (1) |
| 1, Disagree | 0.9% (1) | 0.0% (0) | 1.8% (1) |
| 2, Agree | 30.1% (34) | 31.6% (18) | 29.0% (16) |
| 3, Strongly agree | 64.6% (73) | 61.4% (35) | 68.0% (38) |
| 2. I believe that the health education provided at Pregnancy Pop-Up Village is trustworthy. |  |  |  |
| 0, Strongly disagree | 3.5% (4) | 5.3% (3) | 1.8% (1) |
| 1, Disagree | 4.4% (5) | 3.5% (2) | 5.4% (3) |
| 2, Agree | 29.2% (33) | 36.8% (21) | 21.0% (12) |
| 3, Strongly agree | 62.8% (71) | 54.4% (31) | 71.0% (40) |
| 3. The service providers at Pregnancy Pop-Up Village valued my time. |  |  |  |
| 0, Strongly disagree | 1.8% (2) | 1.8% (1) | 1.8% (1) |
| 1, Disagree | 0.9% (1) | 1.8% (1) | 0.0% (0) |
| 2, Agree | 31.0% (35) | 36.8% (21) | 25.0% (14) |
| 3, Strongly agree | 66.4% (75) | 59.6% (34) | 73.2% (41) |
| 4. The service providers at Pregnancy Pop-Up Village made me feel understood. |  |  |  |
| 0, Strongly disagree | 1.8% (2) | 1.8% (1) | 1.8% (1) |
| 1, Disagree | 0.9% (1) | 1.8% (1) | 0.0% (0) |
| 2, Agree | 30.1% (34) | 33.3% (19) | 26.8% (15) |
| 3, Strongly agree | 67.3% (76) | 63.2% (36) | 71.4% (40) |
| 5. The service providers at Pregnancy Pop-Up Village had my best interests in mind. |  |  |  |
| 0, Strongly disagree | 1.8% (2) | 1.8% (1) | 1.8% (1) |
| 1, Disagree | 2.7% (3) | 3.5% (2) | 1.8% (1) |
| 2, Agree | 33.6% (38) | 38.6% (22) | 29.0% (16) |
| 3, Strongly agree | 61.9% (70) | 56.1% (32) | 68.0% (38) |
| 6. I trust the service providers at Pregnancy Pop-Up Village. |  |  |  |
| 0, Strongly disagree | 1.8% (2) | 1.8% (1) | 1.8% (1) |
| 1, Disagree | 0.0% (0) | 0.0% (0) | 0.0% (0) |
| 2, Agree | 32.7% (37) | 38.6% (22) | 26.8% (15) |
| 3, Strongly agree | 65.5% (74) | 59.6% (34) | 71.4% (40) |
| 7. The service providers at Pregnancy Pop-Up Village helped me feel healthier and well. |  |  |  |
| 0, Strongly disagree | 1.8% (2) | 1.8% (1) | 1.8% (1) |
| 1, Disagree | 1.8% (2) | 1.8% (1) | 1.8% (1) |
| 2, Agree | 36.3% (41) | 43.9% (25) | 28.6% (16) |
| 3, Strongly agree | 60.2% (68) | 52.6% (30) | 67.9% (38) |

**Additional file 2. Bivariate subgroup analysis of sociodemographic characteristics, obstetric history, and care discrimination experiences on the trust score**

|  | **Pregnant/Postpartum (N =57)** | | | | | | **Family (N = 56)** | | | | | |
| --- | --- | --- | --- | --- | --- | --- | --- | --- | --- | --- | --- | --- |
| **Predictor variables** | **Cross Tabs** | | **OLS Regression (Unadjusted)** | | | | **Cross Tabs** | | **OLS Regression (Unadjusted)** | | | |
|  | Mean | SD | Coeff. | *[95% CI]* | | *P-value* | Mean | SD | Coeff. | *[95% CI]* | | *P-value* |
| *Age* |  |  |  |  |  |  |  |  |  |  |  |  |
| 15 - 24 [Reference Group] | 92.4 | 11.9 | 0.0 |  |  |  | 81.0 | 17.2 | 0.0 |  |  |  |
| 25 -34 | 84.0 | 14.2 | -8.4 | -24.8 | 8.0 | 0.159 | 72.0 | 16.4 | -9.0 | -43.6 | 25.6 | 0.469 |
| 35 - 44 | 77.7 | 24.9 | -14.7 | -32.7 | 3.3 | 0.072 | 76.2 | 39.5 | -4.8 | -13.3 | 3.8 | 0.175 |
| 45 and older | 85.7 | 0.0 | -6.7 | . | . | . | 94.9 | 10.8 | 14.0 | -5.2 | 33.1 | 0.103 |
| Unknown | 84.8 | 16.6 | -7.6 | -56.3 | 41.1 | 0.570 | 94.2 | 12.1 | 13.2 | -8.6 | 35.1 | 0.149 |
| *Gender* |  |  |  |  |  |  |  |  |  |  |  |  |
| Female [Reference Group] | 82.2 | 18.9 | 0.0 |  |  |  | 88.9 | 19.3 | 0.0 |  |  |  |
| Male | .. | .. | .. |  |  |  | 81.7 | 20.3 | -7.1 | -21.1 | 6.8 | 0.201 |
| Other/unknown/prefer not to answer | 96.2 | 4.0 | 14.0 | 6.9 | 21.1 | 0.014 | 95.2 | 6.7 | 6.3 | -9.2 | 21.9 | 0.284 |
| *Race/ethnicity* |  |  |  |  |  |  |  |  |  |  |  |  |
| Non-Hispanic Black [Reference Group] | 85.7 | 15.1 | 0.0 |  |  |  | 94.2 | 11.9 | 0.0 |  |  |  |
| Hispanic/Latine | 81.1 | 22.7 | -4.6 | -17.6 | 8.4 | 0.268 | 75.9 | 26.8 | -18.3 | -27.2 | -9.4 | 0.007 |
| Multiracial | 82.7 | 14.2 | -3.0 | -13.3 | 7.2 | 0.331 | 85.0 | 17.3 | -9.2 | -20.6 | 2.3 | 0.084 |
| Other/unknown/prefer not to answer | 92.9 | 4.8 | 7.1 | 7.1 | 7.1 | 0.000 | 100.0 | 0.0 | 5.8 | -5.6 | 17.3 | 0.205 |
| *Language* |  |  |  |  |  |  |  |  |  |  |  |  |
| English [Reference Group] | 87.1 | 14.0 | 0.0 |  |  |  | 92.7 | 13.3 | 0.0 |  |  |  |
| Spanish | 76.8 | 23.1 | -10.3 | -17.6 | -3.0 | 0.026 | 74.4 | 28.5 | -18.3 | -28.6 | -8.0 | 0.011 |
| Other/unknown/prefer not to answer | 90.5 | 13.5 | 3.3 | -35.9 | 42.6 | 0.750 | 91.7 | 8.1 | -1.0 | -7.7 | 5.7 | 0.666 |
| *English proficiency* |  |  |  |  |  |  |  |  |  |  |  |  |
| Very well or well [Reference Group] | 87.5 | 13.8 | 0.0 |  |  |  | 91.7 | 14.0 | 0.0 |  |  |  |
| With difficulty | 73.6 | 29.7 | -13.9 | -44.2 | 16.4 | 0.187 | 77.2 | 33.1 | -14.5 | -24.1 | -4.9 | 0.017 |
| Unknown/prefer not to answer | 74.6 | 12.7 | -12.9 | -21.2 | -4.6 | 0.022 | 80.0 | 18.3 | -11.7 | -23.3 | -0.2 | 0.048 |
| *Number of births* |  |  |  |  |  |  |  |  |  |  |  |  |
| 1-2 births [Reference group] | 85.0 | 13.8 | 0.0 |  |  |  | 91.1 | 14.1 | 0.0 |  |  |  |
| No births | 91.4 | 12.5 | 6.4 | -18.6 | 31.5 | 0.385 | 94.8 | 11.8 | 3.71 | -12.5 | 19.9 | 0.519 |
| 3 or more births | 71.1 | 26.0 | -13.9 | -36.8 | 9.0 | 0.120 | 80.6 | 28.2 | -10 | -21.6 | 0.64 | 0.058 |
| Unknown/prefer not to answer/not applicable | 95.2 | 0.0 | 10.2 | 4.5 | 15.9 | 0.016 | 85.1 | 18.4 | -6 | -24 | 12 | 0.368 |
| *Prenatal care attendance* |  |  |  |  |  |  |  |  |  |  |  |  |
| Yes [Reference Group] | 83.5 | 19.1 | 0.0 |  |  |  | … | … |  |  |  |  |
| No | 91.7 | 16.7 | 8.1 | -27.8 | 44.0 | 0.432 | … | … | … | … | … | … |
| Unknown/prefer not to answer | 76.2 | 13.5 | -7.3 | -23.6 | 8.9 | 0.191 | … | … | … | … | … | … |
| *History of preterm birth* |  |  |  |  |  |  |  |  |  |  |  |  |
| No preterm births [Reference Group] | 84.9 | 18.9 | 0.0 |  |  |  | 93.1 | 13.1 | 0.0 |  |  |  |
| At least 1 preterm birth | 75.7 | 16.9 | -9.2 | -34.3 | 15.8 | 0.254 | 78.6 | 27.4 | -14.6 | -24.8 | -4.4 | 0.02 |
| Unknown | 85.7 | 13.5 | 0.8 | -9.2 | 10.9 | 0.757 | 85.1 | 18.4 | -8.0 | -22.8 | 6.7 | 0.182 |
| *History of pregnancy loss* |  |  |  |  |  |  |  |  |  |  |  |  |
| No prior pregnancy loss [Reference Group] | 87.0 | 14.0 | 0.0 |  |  |  | 89.4 | 20.5 | 0.0 |  |  |  |
| Pregnancy loss | 76.4 | 23.7 | -10.6 | -30.1 | 9.0 | 0.145 | 90.5 | 14.3 | 1.1 | -15.4 | 17.5 | 0.851 |
| Unknown/prefer not to answer/not applicable | 89.3 | 15.2 | 2.3 | -22.4 | 26.9 | 0.730 | 83.1 | 18.0 | -6.3 | -24.5 | 11.9 | 0.352 |
| *Education* |  |  |  |  |  |  |  |  |  |  |  |  |
| High school graduate, GED, or equivalent [Reference Group] | 80.7 | 13.6 | 0.0 |  |  |  | 85.3 | 16.2 | 0.0 |  |  |  |
| Less than high school degree | 80.4 | 32.8 | -0.2 | -48.6 | 48.1 | 0.984 | 82.9 | 17.6 | -2.4 | -27.8 | 23.0 | 0.782 |
| Some college, junior college, or vocational school | 87.7 | 14.4 | 7.0 | -36.3 | 50.3 | 0.557 | 96.2 | 10.3 | 10.8 | -10.0 | 31.7 | 0.197 |
| College graduate and professional or graduate school | 85.3 | 18.1 | 4.6 | -10.6 | 19.9 | 0.320 | 81.5 | 35.0 | -3.8 | -31.7 | 24.1 | 0.694 |
| Unknown/prefer not to answer | 83.7 | 14.8 | 3.0 | -2.3 | 8.3 | 0.136 | 88.9 | 19.2 | 3.5 | -5.5 | 12.6 | 0.302 |
| *Employment status* |  |  |  |  |  |  |  |  |  |  |  |  |
| Unemployed [Reference Group] | 81.1 | 20.4 | 0.0 |  |  |  | 90.5 | 14.4 | 0.0 |  |  |  |
| Full-time | 85.7 | 13.5 | 4.6 | -5.8 | 15.1 | 0.198 | 82.9 | 19.8 | -7.6 | -16.9 | 1.6 | 0.079 |
| Part-time | 93.7 | 11.4 | 12.6 | -12.9 | 38.0 | 0.168 | 55.6 | 50.9 | -34.9 | -109.8 | 39.9 | 0.234 |
| Unknown/prefer not to answer | 75.2 | 21.9 | -5.9 | -18.2 | 6.5 | 0.177 | 94.4 | 13.6 | 4.0 | -9.4 | 17.3 | 0.414 |
| *Income assistance* |  |  |  |  |  |  |  |  |  |  |  |  |
| No [Reference Group] | 87.2 | 12.8 | 0.0 |  |  |  | 90.5 | 14.9 | 0.0 |  |  |  |
| Yes | 80.8 | 21.8 | -6.4 | -30.4 | 17.6 | 0.369 | 85.1 | 23.6 | -5.4 | -23.9 | 13.2 | 0.425 |
| Unknown/ prefer not to answer | 85.0 | 15.9 | -2.2 | -14.5 | 10.1 | 0.525 | 100.0 | 0.0 | 9.5 | 0.2 | 18.9 | 0.048 |
| *Residence* |  |  |  |  |  |  |  |  |  |  |  |  |
| Bayview-Hunter's Point (San Francisco) [Reference Group] | 80.7 | 23.4 | 0.0 |  |  |  | 93.0 | 13.4 | 0.0 |  |  |  |
| Other San Francisco | 86.3 | 15.1 | 5.6 | -14.5 | 25.6 | 0.356 | 85.9 | 23.3 | -7.1 | -31.7 | 17.5 | 0.426 |
| East Bay | 80.3 | 13.6 | -0.5 | -20.5 | 19.6 | 0.930 | 76.2 | 11.7 | -16.8 | -25.6 | -8.0 | 0.009 |
| Other/unknown/prefer not to answer | 87.3 | 18.0 | 6.6 | -44.3 | 57.5 | 0.635 | 92.9 | 13.5 | -0.1 | -16.7 | 16.5 | 0.980 |
| *Housing status* |  |  |  |  |  |  |  |  |  |  |  |  |
| Rent home or apartment [Reference Group] | 81.1 | 20.5 | 0.0 |  |  |  | 88.3 | 23.1 | 0.0 |  |  |  |
| Homeless shelter | 85.0 | 16.1 | 3.9 | -21.5 | 29.3 | 0.575 | 76.2 | 14.7 | -12.1 | -38.7 | 14.5 | 0.244 |
| Owns home or apartment | 87.3 | 18.0 | 6.2 | -10.7 | 23.1 | 0.256 | 93.3 | 9.3 | 5.1 | -17.7 | 27.8 | 0.529 |
| Public housing | 80.3 | 22.2 | -0.8 | -54.3 | 52.6 | 0.952 | 90.0 | 16.1 | 1.7 | -20.3 | 23.7 | 0.818 |
| Unknown/Other (e.g., living with someone for free, no living place, transitional housing etc.) | 90.5 | 11.0 | 9.4 | -13.2 | 31.9 | 0.216 | 91.1 | 12.8 | 2.8 | -16.7 | 22.3 | 0.678 |
| *Social Support* |  |  |  |  |  |  |  |  |  |  |  |  |
| Yes, definitely [Reference Group] | 87.7 | 13.3 | 0.0 |  |  |  | 91.2 | 20.2 | 0.0 |  |  |  |
| No, not at all | 77.4 | 15.7 | -10.3 | -28.6 | 7.9 | 0.135 | 84.0 | 16.0 | -7.2 | -22.4 | 7.9 | 0.226 |
| A little | 80.4 | 20.9 | -7.4 | -19.1 | 4.4 | 0.114 | 85.7 | 17.2 | -5.5 | -16.6 | 5.6 | 0.212 |
| Somewhat | 76.2 | 26.7 | -11.5 | -68.0 | 45.0 | 0.473 | 83.1 | 20.4 | -8.2 | -20.9 | 4.6 | 0.134 |
| Unknown/prefer not to answer | 95.2 | 0.0 | 7.5 | -5.9 | 20.9 | 0.137 | 100.0 | 0.0 | 8.8 | -7.1 | 24.7 | 0.177 |
| *Medical Insurance status* |  |  |  |  |  |  |  |  |  |  |  |  |
| Public insurance (e.g., Medicaid, Medi-Cal, etc.) [Reference Group] | 86.0 | 19.4 | 0.0 |  |  |  | 90.0 | 19.4 | 0.0 |  |  |  |
| Private or employer provided insurance | 82.7 | 13.7 | -3.3 | -19.3 | 12.8 | 0.471 | 90.5 | 16.3 | 0.5 | -13.1 | 14.0 | 0.916 |
| No insurance | 66.7 | 0.0 | -19.3 | -35.6 | -3.0 | 0.036 | 78.6 | 20.0 | -11.4 | -25.8 | 3.0 | 0.086 |
| Unknown/prefer not to answer | 73.5 | 19.4 | -12.5 | -57.2 | 32.2 | 0.352 | 81.7 | 20.3 | -8.2 | -20.8 | 4.3 | 0.128 |
| *Food insecurity (Worried food would run out)* | | |  |  |  |  |  |  |  |  |  |  |
| Never true [Reference Group] | 90.0 | 12.6 | 0.0 |  |  |  | 94.6 | 11.6 | 0.0 |  |  |  |
| Sometimes true | 85.5 | 13.0 | -4.6 | -22.4 | 13.3 | 0.386 | 69.8 | 30.5 | -24.7 | -32.6 | -16.8 | 0.002 |
| Often true | 63.8 | 38.3 | -26.2 | -74.0 | 21.6 | 0.142 | 82.7 | 17.3 | -11.8 | -22.0 | -1.6 | 0.035 |
| Unknown/prefer not to answer | 77.0 | 18.4 | -13.0 | -33.0 | 6.9 | 0.106 | 86.9 | 20.3 | -7.7 | -17.4 | 2.1 | 0.089 |
| *Food insecurity (Insufficient funds)* |  |  |  |  |  |  |  |  |  |  |  |  |
| Never true [Reference Group] | 86.7 | 16.2 | 0.0 |  |  |  | 94.6 | 11.7 | 0.0 |  |  |  |
| Sometimes true | 85.9 | 13.9 | -0.8 | -28.9 | 27.3 | 0.916 | 73.2 | 29.5 | -21.5 | -39.6 | -3.4 | 0.033 |
| Often true | 68.0 | 32.8 | -18.7 | -70.4 | 33.0 | 0.26 | 82.5 | 17.5 | -12.1 | -22.0 | -2.2 | 0.03 |
| Unknown/prefer not to answer | 82.9 | 15.4 | -3.9 | -29.5 | 21.8 | 0.584 | 89.3 | 15.4 | -5.3 | -20.3 | 9.7 | 0.34 |
| *Relationship status* |  |  |  |  |  |  |  |  |  |  |  |  |
| Married/partnered, living together [Reference Group] | 82.9 | 22.0 | 0.0 |  |  |  | 79.9 | 16.9 | 0.0 |  |  |  |
| Married/partnered, not living together | 79.5 | 13.7 | -3.4 | -24.8 | 18.0 | 0.564 | 84.9 | 16.9 | 5.1 | 2.7 | 7.4 | 0.006 |
| Single | 86.4 | 14.5 | 3.5 | -22.8 | 29.7 | 0.628 | 91.4 | 20.0 | 11.6 | -3.5 | 26.7 | 0.093 |
| Other/unknown/prefer not to answer | 84.7 | 20.7 | 1.7 | -33.5 | 36.9 | 0.853 | 100.0 | 0.0 | 20.1 | 16.8 | 23.5 | 0.000 |
| *Everyday discrimination experience* |  |  |  |  |  |  |  |  |  |  |  |  |
| Never [Reference Group] | 86.1 | 14.8 | 0.0 |  |  |  | 84.1 | 19.0 | 0.0 |  |  |  |
| Rarely | 83.0 | 25.6 | -3.1 | -44.0 | 37.8 | 0.774 | 86.4 | 27.3 | 2.3 | -11.9 | 16.4 | 0.645 |
| Sometimes | 81.4 | 17.9 | -4.7 | -15.9 | 6.4 | 0.209 | 90.7 | 14.8 | 6.6 | -14.2 | 27.3 | 0.388 |
| Often | 85.7 | 8.9 | -0.4 | -8.8 | 8.0 | 0.845 | 89.5 | 15.8 | 5.4 | -15.6 | 26.4 | 0.474 |
| *Prenatal care discrimination experience* | |  |  |  |  |  |  |  |  |  |  |  |
| Never [Reference Group] | 82.5 | 24.7 | 0.0 |  |  |  | 90.0 | 20.9 | 0.0 |  |  |  |
| Rarely | 83.8 | 17.4 | 1.3 | -38.2 | 40.7 | 0.903 | 86.4 | 15.2 | -3.6 | -12.1 | 4.9 | 0.267 |
| Sometimes | 82.5 | 14.5 | 0.0 | -39.5 | 39.5 | 1.000 | 83.9 | 18.7 | -6.1 | -24.6 | 12.3 | 0.367 |
| Often | 93.7 | 7.3 | 11.1 | -21.2 | 43.5 | 0.277 | 92.4 | 14.5 | 2.4 | -15.9 | 20.7 | 0.708 |

Abbreviations: 95% CI: 95% confidence interval; SD: standard deviation
